# Mental health and educational attainment: Replicating diminishing associations in an England cohort

**DOI:** 10.64898/2026.03.20.26348881

**Authors:** Alice Wickersham, Emma Soneson, Nicoletta Adamo, Craig Colling, Amelia Jewell, Johnny Downs

## Abstract

**Background:** A study conducted in Norway showed that the association between pupil mental health diagnoses and educational attainment has weakened over time. One possible explanation is that earlier detection of mental health problems in recent years has facilitated earlier treatment, intervention, and educational support that might improve academic outcomes. We investigated whether the weakening association between mental health and attainment could be replicated in England, and explained by earlier age at first diagnosis.

**Methods:** This was a secondary longitudinal data analysis of de-identified records from a secondary mental healthcare provider in England, which have been linked to the Department for Education’s National Pupil Database. We included n=149,841 pupils residing in South East London, born 1993-2003, who completed their end-of-school exams 2009-2019. The main exposure variables were ADHD and internalising disorder diagnosis. In linear regressions, we investigated their associations with Year 11 attainment (typically assessed age 15-16 years), whether this was modified by birth year, and the role of age at first diagnosis.

**Results:** On average, ADHD (n=844, 0.6%) and internalising disorder (n=2,523, 1.7%) were associated with lower Year 11 attainment. However, significant interactions between diagnosis and birth year suggested that pupils with these disorders showed increases in standardised exam scores over successive birth cohorts, resulting in a closing attainment gap over time. While age at first diagnosis became younger over the period, this did not confound the observed associations.

**Conclusions:** We replicated findings from Norway that suggest a narrowing attainment gap between those with and without ADHD and internalising disorder diagnoses. Building on this, we ruled out earlier age of diagnosis as a possible explanation for this phenomenon. With administrative data research growing internationally, we are increasingly able to replicate mental health and education trends in different countries, opening more opportunities for international collaboration.

## Introduction

Recent years have shown a rise in the prevalence of mental health problems among young people, with most recent estimates for England showing that one in five children and young people have a probable mental health disorder (Newlove-Delgado et al., 2023). These young people can experience a range of difficulties, including in their educational outcomes. For example, research has shown that children and adolescents with depression have lower school attendance than their peers (Finning et al., 2019) and are less likely to perform at expected levels in end-of-school exams (Dalsgaard et al., 2020; Wickersham et al., 2023; Wickersham et al., 2020; Wickersham, Dickson, Stewart, Ford, & Downs, 2019). Concerningly, analyses using UK cohort and epidemiological survey data suggest that the impacts of child and adolescent mental health problems, including on school functioning and academic attainment, have increased in recent decades (Armitage, Newlove-Delgado, Ford, McManus, & Collishaw, 2025; Sellers et al., 2019).

However, recent evidence from Norway contradicts these findings by suggesting that the attainment gap for those with diagnosed mental health disorders may be becoming less pronounced (Nordmo et al., 2024). Nordmo and colleagues’ (2024) analysis of population-wide register data showed that, while the lifetime prevalences of internalising disorder and ADHD diagnoses received from primary care have increased since 2006, the strength of their association with educational attainment has diminished over time. For diagnoses recorded in specialist care data, there was no clear trend in prevalence, but again, the association between diagnosed disorders and educational attainment weakened over time. The authors proposed various possible explanations for these findings. For example, the threshold for receiving a mental health diagnosis may have decreased, such that individuals with less severe symptoms and functional impairment are increasingly likely to be diagnosed. They also considered that improving treatment coverage or lessening stigma could be mitigating the negative impact that disorders have on educational outcomes.

Another as-yet unexplored explanation could be found in age at diagnosis. Over successive years since the 1970s, mental health disorders have been first diagnosed at progressively younger ages (Plana-Ripoll et al., 2022). Earlier detection and diagnosis of mental health problems could plausibly allow more time to implement effective treatment strategies before end-of-school exams, thus mitigating the long-term impact of these disorders across many domains, including attainment outcomes (Anderson et al., 2019; McGorry & Mei, 2018). In addition to facilitating earlier access to formal mental health care, earlier diagnoses could also enable schools to implement interventions or adjustments that could enhance pupils’ school functioning and support academic performance (DuPaul et al., 2021; Moore et al., 2018).

In this study, we explored this phenomenon further using linked health and education records in a cohort from South London, England. First, we aimed to reproduce the methods used in Nordmo et al.’s (2024) study, not only to investigate whether their findings on diminishing associations between mental health disorders and attainment generalise beyond Norway, but also to demonstrate the feasibility of replicating administrative data linkage studies internationally. Second, we aimed to extend this analysis by investigating our hypothesis that these associations may be explained, in part, by earlier age at first diagnosis.

## Methods

### Design and sample

Reporting follows the STROBE checklist for cohort studies (Table S1). We conducted a retrospective cohort study using an existing data linkage between South London and Maudsley NHS Foundation Trust (SLaM) and the Department for Education (DfE). Full details of this individual-level linkage can be found elsewhere (Downs et al., 2019). Briefly, anonymised electronic health records for children and adolescents referred to SLaM CAMHS are accessible for research via the Clinical Record Interactive Search (CRIS) (Perera et al., 2016). These records were linked to DfE’s National Pupil Database (NPD), which contains education records for all pupils in England’s state-maintained schools. The linkage was most recently refreshed up to the 2018/19 academic year.

In this study, we included all pupils born 1993 to 2003, who resided in the SLaM catchment area (Croydon, Southwark, Lewisham, Lambeth), were captured in the NPD’s Key Stage 4 data table, and were age 15 at the start of their Key Stage 4 academic year. These criteria were designed to emulate Nordmo et al.’s inclusion criteria as closely as possible; in that study, authors included birth years 1990 to 2003 for primary care analyses and 1992 to 2003 for specialist care analyses. In total, n=150,126 were eligible for inclusion.

### Outcome

End-of-school exam performance was derived from the NPD’s Key Stage 4 data table. In England, Key Stage 4 is typically assessed in Year 11 of secondary school, at age 15/16 years. Assessments comprise General Certificate of Secondary Education (GCSE) exams, and other equivalents. We used capped point scores on these assessments, which is based on each pupil’s best eight GCSE or equivalent grades. These were standardised as z-scores within each academic year to account for changes in scoring over time. For pupils who had duplicate records (n = 196), we kept their earliest attempt (i.e. their score in the first academic year in which the exam was taken); for those with multiple attempts within one year, we kept their record with the highest point score. Henceforth we use the term ‘Year 11 attainment’ for this variable.

### Exposures

*Birth year* was derived from the NPD’s Key Stage 4 data table. *ADHD* and *internalising disorder* were derived from primary and secondary diagnosis fields in CRIS, first diagnosed any time before the end (31 August) of Year 11 (the assessment year). In accordance with Nordmo et al., ADHD was defined using ICD-10 codes F90.0 and F90.1, and internalising disorder was defined using ICD-10 codes F32.1, F33.1, F32.0, F33.0, F34.1, F40.1, F41.2, F41.0, F41.1, F41.9, and F43.1 (capturing depressive disorders, anxiety disorders, and post-traumatic stress disorder). *Age at first diagnosis* was calculated using the earliest diagnosis date from CRIS for these ICD-10 codes, and month and year of birth from the NPD’s Key Stage 4 data table. Finally, *gender* was derived from the NPD’s Key Stage 4 data table, where it is recorded as a binary variable (male, female).

### Statistical analysis

We used descriptive statistics to explore and plot changing age at first diagnosis and attainment in each disorder group over time. For all statistical models, ADHD and internalising disorder were examined in separate regressions, consistent with Nordmo et al. To address our first aim, we conducted linear regressions with Year 11 attainment z-score as the outcome. First, we included birth year and disorder status as the exposure variables. Then we tested for an interaction between birth year and disorder status using a Wald test.

Significant interactions were further explored with linear regressions stratified by diagnosis. To address our second aim, we limited the sample to those with each disorder, and in separate models for each disorder we repeated linear regressions with Year 11 attainment z-score as the outcome, and birth year and age at first diagnosis as exposures. All models adjusted for gender as a covariate.

Of the eligible sample, n=285 (0.2%) were missing data on Year 11 attainment. Given that this affected a small proportion of the eligible sample and the remaining pupils had no missing data on the exposure variables, we conducted complete case analyses (n=149,841). The significance level was set at p<0.05. All analyses were conducted in Stata version 17.0; visualisations were made in R version 4.2.1.

## Results

Of the n=149,841 pupils analysed, 0.6% received an ADHD diagnosis, and 1.7% received an internalising disorder diagnosis (Table 1). The prevalence of recorded ADHD diagnoses increased from 0.1% among those born in 1993, to 1.1% among those born in 2003. The prevalence of recorded internalising disorder diagnoses increased from 0.7% to 1.9% in the same birth cohorts (Figure S1).

**Table 1:** Sample descriptive statistics, n=149,841.

| Variable | Descriptive statistics |
| --- | --- |
| Gender, n (%) |  |
| Male | 75,699 (50.5%) |
| Female | 74,142 (49.5%) |
| ADHD diagnosis, n (%) | 844 (0.6%) |
| Age at first ADHD diagnosis, mean (SD) | 11.8 (2.7) |
| Internalising disorder diagnosis, n (%) | 2,523 (1.7%) |
| Age at first internalising disorder diagnosis, mean (SD) | 13.8 (2.0) |
| Year 11 exam z-score, mean (SD) | 0.00 (1.00) |

Linear regression analyses showed that, on average, pupils who received an ADHD or internalising disorder diagnosis had lower Year 11 attainment than those who did not (Table 2). Birth year significantly modified the association between ADHD diagnosis and Year 11 attainment (Wald test p<0.001; Table S1), and between internalising disorder diagnosis and Year 11 attainment (Wald test p<0.001; Table S2). Stratified linear regressions showed that birth year was positively associated with Year 11 attainment among those with these disorders, but not those without (Table 3), indicating a closing attainment gap between pupils with and without these disorders (Figure 1).

**Table 2:**
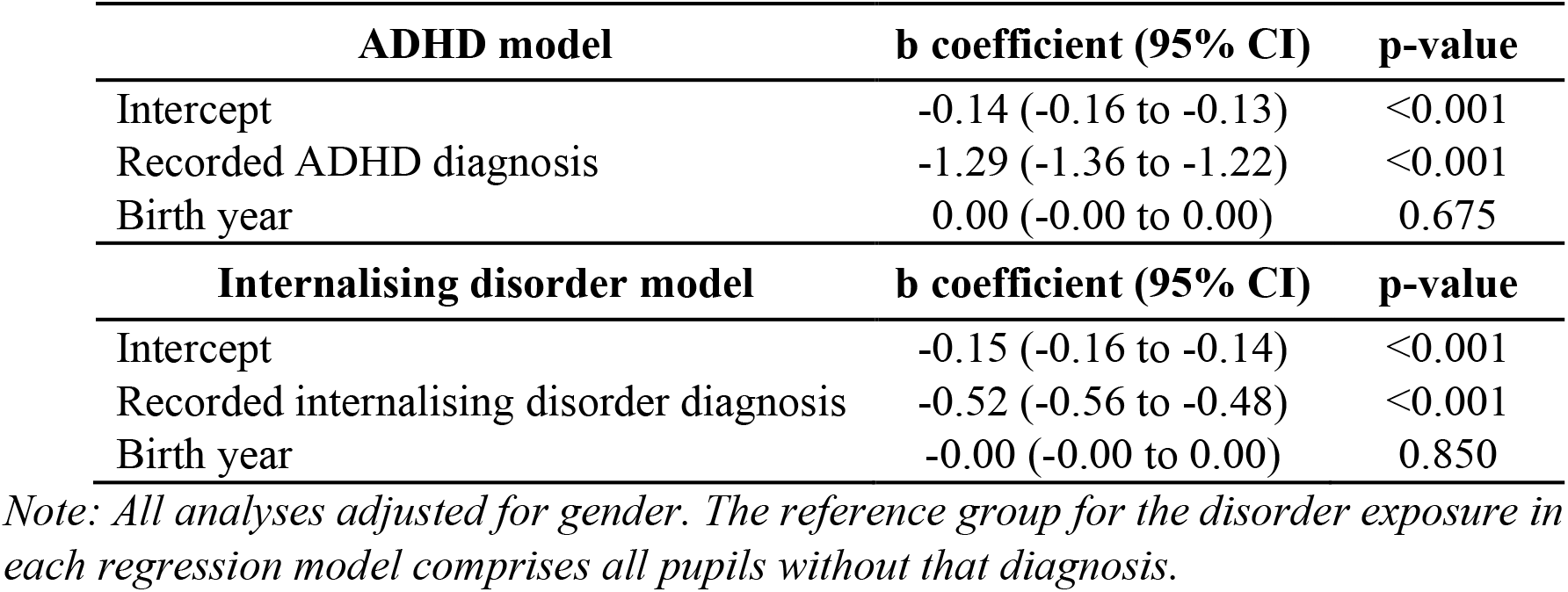
Linear regressions between mental health, birth year, and Year 11 attainment z-score (n=149,841).

**Table 3:** Stratified linear regressions between birth year and Year 11 attainment z-score.

| **ADHD stratified models** | **b coefficient (95% CI)** | **p-value** |
| --- | --- | --- |
| **No recorded ADHD diagnosis (n=148,997)** |  |  |
| Intercept | -0.14 (-0.15 to -0.13) | <0.001 |
| Birth year | 0.00 (-0.00 to 0.00) | 0.901 |
| **Recorded ADHD diagnosis (n=844)** |  |  |
| Intercept | -1.81 (-1.99 to -1.62) | <0.001 |
| Birth year | 0.06 (0.03 to 0.09) | <0.001 |
| **Internalising disorder stratified models** | **b coefficient (95% CI)** | **p-value** |
| **No recorded internalising disorder diagnosis (n=147,318)** |  |  |
| Intercept | -0.14 (-0.15 to -0.13) | <0.001 |
| Birth year | -0.00 (-0.00 to 0.00) | 0.319 |
| **Recorded internalising disorder diagnosis (n=2,523)** |  |  |
| Intercept | -1.07 (-1.19 to -0.95) | <0.001 |
| Birth year | 0.05 (0.03 to 0.07) | <0.001 |

*Note: All analyses adjusted for gender.*

**Figure 1.**
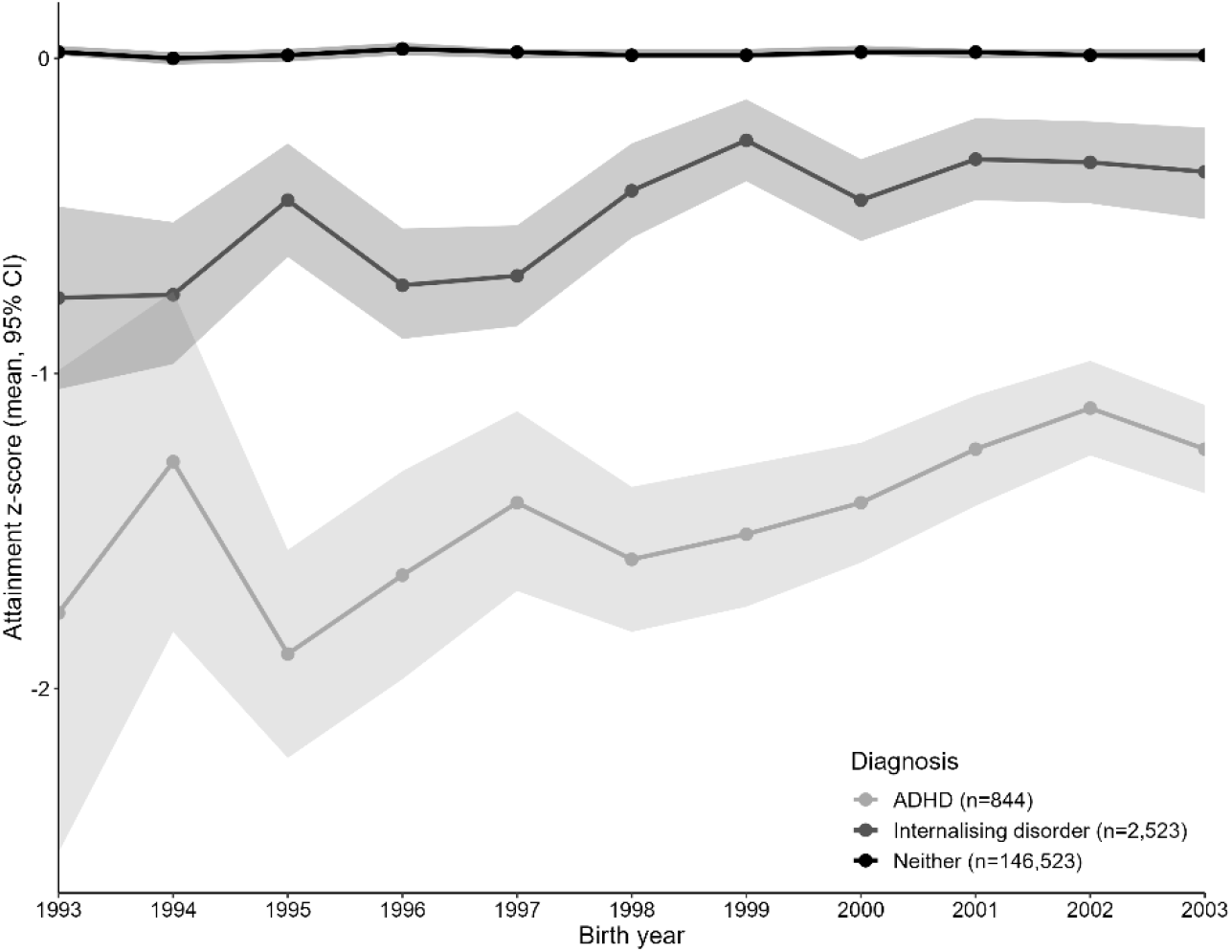
Average Year 11 attainment for each birth year, stratified by mental health diagnosis.

For both ADHD and internalising disorder, the average age at first diagnosis declined over time (Figure 2). Limiting the sample to pupils with each disorder, we repeated linear regressions between birth year and Year 11 attainment while adjusting for age at first diagnosis. Age at first diagnosis did not confound the association between birth year and Year 11 attainment for either disorder (Table 4), signalling that earlier diagnoses have not contributed to the narrowing attainment gap.

**Table 4:** Linear regressions between birth year and Year 11 attainment z-score, adjusted for age at first diagnosis.

| ADHD model (n=844) | b coefficient (95% CI) | p-value |
| --- | --- | --- |
| Intercept | -1.53 (-1.96 to -1.10) | <0.001 |
| Birth year | 0.05 (0.02 to 0.08) | <0.001 |
| Age at first diagnosis | -0.02 (-0.05 to 0.01) | 0.156 |
| Internalising disorder model (n=2,523) | b coefficient (95% CI) | p-value |
| Intercept | -1.04 (-1.40 to -0.68) | <0.001 |
| Birth year | 0.05 (0.03 to 0.07) | <0.001 |
| Age at first diagnosis | -0.00 (-0.03 to 0.02) | 0.867 |

Note: All analyses adjusted for gender.

**Figure 2.**
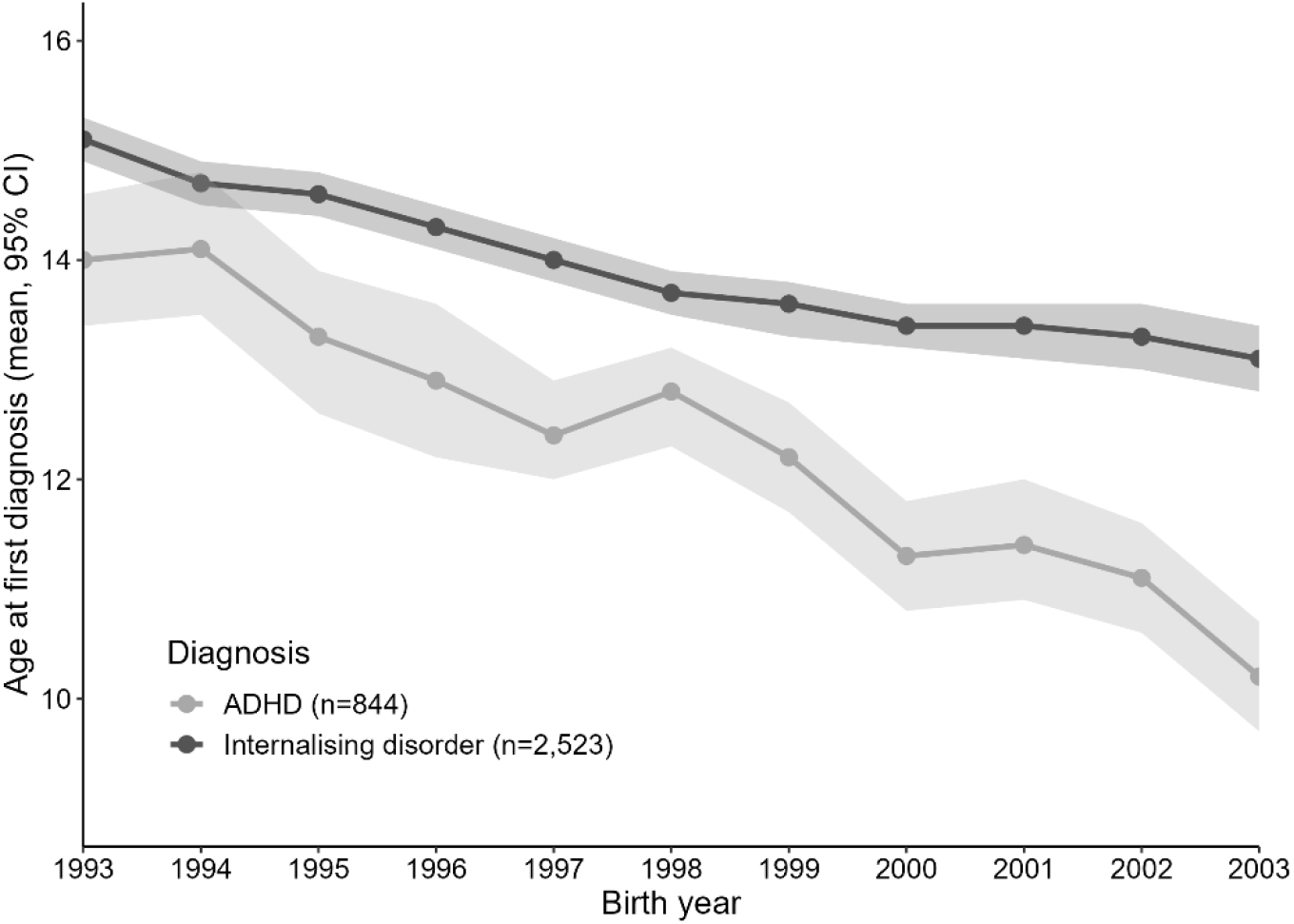
Age at first mental health diagnosis for each birth cohort.

### Post-hoc exploratory analysis

In a *post-hoc* exploratory analysis, we investigated whether there was also a closing attainment gap for our cohort at Key Stage 2, typically assessed in Year 6 at age 10/11 years. Due to cell counts <10 in earlier birth years, we limited these analyses to birth years 1997-2003. A total of n=85,055 pupils from these birth years in our original cohort had available Key Stage 2 data, and were age 10 at the start of their Key Stage 2 academic year. Year 6 attainment z-scores were derived from English and Maths performance up to 2011/12, and Maths performance only from 2012/13. These exploratory analyses generally rely on smaller cell sizes, so should be interpreted with caution.

Of this sample, n=258 (0.3%) pupils were first diagnosed with ADHD and n=248 (0.3%) with internalising disorder before 31st August of their Key Stage 2 assessment year. In total, 41.1% of those with ADHD diagnoses before the end of Year 11 already had a diagnosis by the end of Year 6, compared to only 14.5% of those with internalising disorder diagnoses (Table S3 and Table S4).

Linear regression analyses showed that, on average, pupils who received an ADHD or internalising disorder diagnosis before the end of Year 6 had lower attainment than those who did not (Table 5). Birth year did not modify the association between ADHD diagnosis and Year 6 attainment (Wald test p = 0.166; Table S5), but did significantly modify the association between internalising disorder diagnosis and Year 6 attainment (Wald test p=0.007; Table S6). Stratified linear regressions showed that birth year was positively associated with Year 6 attainment among those with internalising disorder diagnoses, but not those without (Table 6), indicating a closing attainment gap between pupils with and without diagnosis.

**Table 5:** Linear regressions between mental health, birth year, and Year 6 attainment z-score (n=85,055).

| **ADHD model** | **b coefficient (95% CI)** | **p-value** |
| --- | --- | --- |
| Intercept | -0.05 (-0.06 to -0.03) | <0.001 |
| Recorded ADHD diagnosis | -0.91 (-1.03 to -0.79) | <0.001 |
| Birth year | -0.00 (-0.01 to 0.00) | 0.210 |
| **Internalising disorder model** | **b coefficient (95% CI)** | **p-value** |
| Intercept | -0.05 (-0.06 to -0.04) | <0.001 |
| Recorded internalising disorder diagnosis | -0.17 (-0.30 to -0.05) | 0.006 |
| Birth year | -0.00 (-0.01 to 0.00) | 0.111 |

*Note: All analyses adjusted for gender. The reference group for the disorder exposure in each regression model comprises all pupils without that diagnosis.*

**Table 6:**
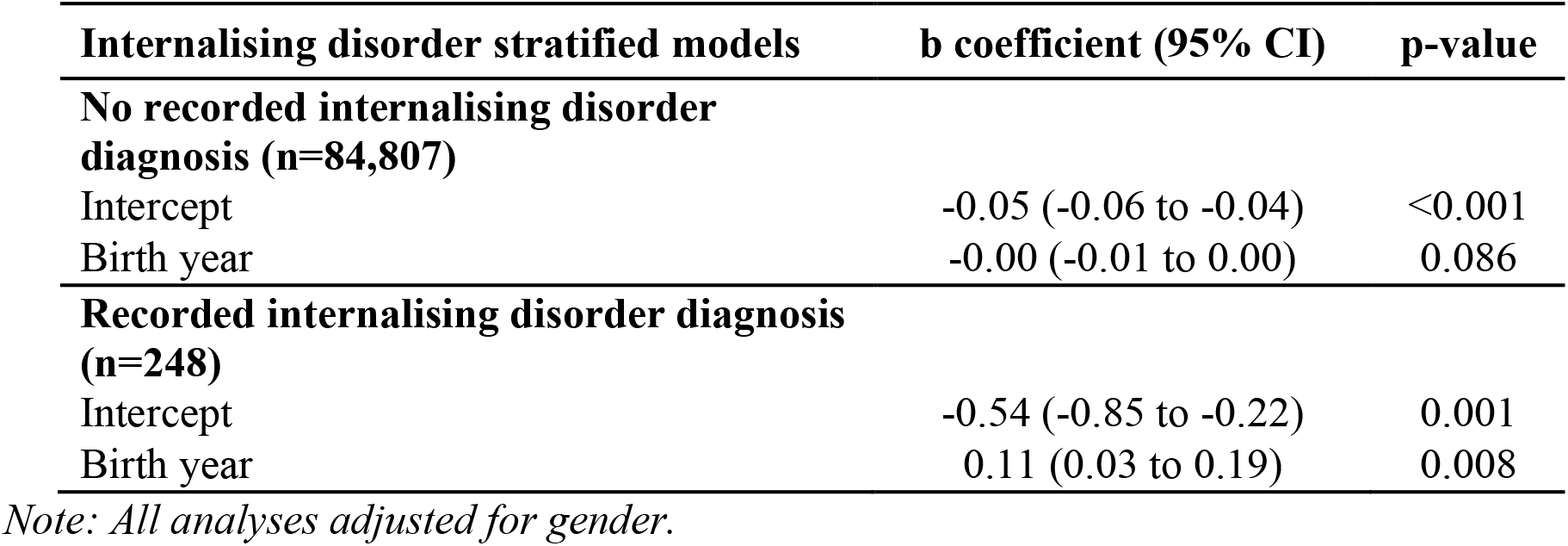
Stratified linear regression between birth year and Year 6 attainment z-score.

## Discussion

In this study, we replicated Nordmo et al.’s (2024) findings that both ADHD and internalising disorder diagnoses showed diminishing associations with performance on end-of-school exams (age 15-16) from 2009 to 2019, indicating a narrowing attainment gap for those with mental health disorders diagnosed in a secondary care setting. Our findings suggest that this is an internationally observable trend. We also showed that, contrary to our second hypothesis, this trend is not explained by the earlier age at first diagnosis seen over recent years. In *post-hoc* exploratory analyses, we found that, for internalising disorder but *not* ADHD diagnoses, there was also evidence within this same cohort of a narrowing attainment gap for exams administered at the end of primary school (age 10-11), offering additional context for understanding these findings.

Overall, our findings suggest that pupils receiving these diagnoses may be experiencing less impairment in their educational outcomes, as compared to previous years. One suite of potential explanations pertains to changes in service capacity, diagnostic criteria, and help-seeking behaviours that have altered the landscape of who receives a diagnosis, such that young people receiving diagnoses in later cohorts have less functional impairment. In theory, it is possible that increased spending and resources in mental health (Children’s Commissioner for England, 2021) could be increasing the capacity of services like CAMHS to diagnose and treat less severe cases (Scott, 2023). However, there is a lack of data to explore whether this might be true. Any potential lowering of thresholds for referral/acceptance to CAMHS are also unlikely to fully explain the findings, as Nordmo et al. (2024) observed the same trend using diagnoses from primary care. Lowering thresholds for diagnosis could also explain the findings, with later cohorts of young people able to receive diagnoses with lower levels of functional impairment. Lessening stigma and increasing awareness of mental health could be leading more individuals to seek diagnoses irrespective of their functional outcomes (Tam, Wu, Zhang, Pawliuk, & Robillard, 2024).

While our findings do not lend support to the hypothesis that earlier age at first diagnosis is allowing more time for successful intervention before end-of-school exams, it remains possible that the narrowing attainment gap reflects more effective mental health interventions which, in turn, have a positive impact on educational outcomes. However, this would not be consistent with literature suggesting that youth treatment benefit has not generally increased over the years (Weisz, Venturo-Conerly, Fitzpatrick, Frederick, & Ng, 2023); indeed, only a minority of young people accessing mental health services seem to show meaningful improvements in symptoms and functioning (Krause, Edbrooke-Childs, Singleton, & Wolpert, 2022).

Yet another potential explanation is that progress and innovation within the education sector may be reducing inequalities in exam performance. The role of schools in mental health promotion, prevention, and early intervention has increased in recent decades (Fazel & Soneson, 2023; Hoover & Bostic, 2021), with growing expectations that schools take an active role in supporting young people with mental health problems (Department of Health and Social Care and Department for Education, 2017). It would be reasonable to suggest that increased awareness and knowledge amongst school staff about the impacts of mental health problems may have contributed to more regular implementation of interventions, tailored strategies, and adjustments to support pupils with mental health problems, targeting either the mental health problems themselves (Gee et al., 2020; Public Health England and Department for Education, 2021) or their impacts on academic functioning. And, whilst such support need not rely on having a diagnosis, formal diagnoses may open the door for additional funding and more formal provision.

To help differentiate these possible explanations, further data could helpfully explore trends of educational experiences preceding diagnosis. Our current data on end-of-school exams leaves it unclear whether pupils already have lower levels of functional impairment at the point of receiving their CAMHS diagnosis, or whether they are merely showing lower levels of functional impairment by the time they sit end-of-school exams. The study by Nordmo et al. (2024) provides some insight in this regard, as their measure of educational performance is a 10-year cumulative grade point average (GPA) comprising both teacher-evaluated grades and externally graded exams. Although more granular longitudinal data would still offer additional opportunities to explore temporal associations, the GPA measure does at least demonstrate that the closing attainment gap extends beyond end-of-school exam performance alone.

In an attempt to explore whether this narrowing gap was evident at earlier assessments, we conducted a *post hoc* analysis of Year 6 attainment. This analysis yielded interesting results and divergent patterns across diagnostic categories. Whilst 41% of pupils who would go on to have an ADHD diagnosis before Year 11 exams already had a diagnosis at these earlier exams, this was only true for 15% of those with internalising diagnoses. This is not unexpected given the distinct developmental trajectories of the different diagnoses (Solmi et al., 2022), but is notable in that the trend for a narrowing attainment gap was apparent only for those with internalising disorder diagnoses at Year 6. Whilst these results merit more systematic investigation, these early findings suggest that the mechanisms behind the attainment gap may be diagnosis-specific. It was also notable that, for pupils with internalising disorder diagnoses, the birth year coefficient was larger in Year 6 than Year 11 (average attainment z-score increase per birth year was +0.11 in Year 6, versus +0.05 in Year 11), suggesting that reasons behind the narrowing attainment gap might be amplified in earlier years. The additional context offered by these findings may therefore suggest potential areas for further investigation.

Another avenue for future research might be exploring the potential effects of in-school support for pupils with mental health diagnoses. Although the lack of systematic documentation of school-based interventions makes this challenging, administrative data do capture information on whether pupils are receiving Special Educational Needs provision. Furthermore, as raised by Nordmo et al. (2024), it would also be helpful to understand whether the attainment gap is narrowing for all sociodemographic groups, particularly according to socioeconomic status. Indeed, it is plausible that the closing gap we observed could be under-estimated by our inability to capture diagnoses that have been sought through private healthcare.

A final area that merits additional exploration lies in the divergence of findings between studies using administrative data, such as ours and Nordmo et al.’s (2024), which suggest a diminishing association between mental health problems and educational outcomes, and those using survey data, which suggest the opposite (Armitage et al., 2025; Sellers et al., 2019). Heterogeneity in data sources, geographical coverage, and conceptualisation and measurement of exposures and outcomes may contribute to these apparently conflicting findings. Linkage of population-based surveys with administrative health and education data offers one potential approach to more fully understand and contextualise our findings (Edwards et al., 2023).

Methodologically, this study highlights the feasibility of replicating mental health and education research internationally. Owing to the increasing availability of administrative data linkages between these areas, there is more opportunity than ever to collaborate and corroborate work across countries (Wickersham & Downs, 2023). This replication work is essential; by identifying overlapping trends, countries can work jointly on finding solutions in full confidence that many of us are grappling with the same challenges, despite having mental health and education systems that diverge operationally. It is particularly noteworthy that findings of the closing attainment gap replicated across Norway and England despite differences in (1) population included (all pupils in the Norwegian study versus only pupils in state-funded education in ours); (2) geographic coverage (national in the Norwegian study and one London mental health trust’s catchment area in ours); (3) differing time trends in the apparent prevalence of specialist diagnosis (inconsistent in the Norwegian study and increasing in ours); and (4) operationalisation of academic attainment (cumulative 10-year GPA in the Norwegian study and end-of-school exam performance in ours). Beyond these observed differences, there will additionally be a range of contextual and cultural differences in the two countries’ health and education systems not captured in our data, making the replicated findings all the more convincing. It is important to note that this sort of work is supported by transparent and thorough reporting by study authors; Nordmo et al.’s (2024) clear and exhaustive methodological reporting enabled us to independently replicate their process, a goal which all researchers should strive for when reporting their studies.

Several limitations of our study should be acknowledged. First, our sample was limited to pupils in state-maintained schools residing in South London, such that its generalisability to the rest of England and the United Kingdom is uncertain. However, given that this study was itself a replication of work conducted in Norway, it is reasonable to consider that these findings may impact multiple regions and developed countries. Second, while we focused on diagnoses received before end-of-school exams, causality between mental health and educational outcomes cannot be established. As outlined above, further work taking into account educational attainment which precedes diagnosis could shed further light on this.

Third, the CRIS-NPD linkage does not capture young people who are not in school, which may bias results as this is a population that has grown over time, and these young people are more likely than their peers to have mental health problems (Children’s Commissioner for England, 2024; Ryder, Edwards, & Rix, 2017). Fourth, it remains possible that observed trends could be impacted by changes in exam score distributions over time, although we standardised attainment within each academic year to make scores as comparable as possible. Finally, our definition of internalising disorders encompassed depressive disorders, anxiety disorders, and post-traumatic stress disorder, derived from structured fields containing ICD-10 codes. We did this to precisely replicate the definitions used by Nordmo et al. (2024); however, given the potentially differential effects of depression and anxiety on attainment (Riglin, Petrides, Frederickson, & Rice, 2014; Socratous et al., 2026), future studies could helpfully explore more granular diagnostic groupings, and could additionally derive diagnosis data from free-text notes using Natural Language Processing techniques.

## Conclusion

Diminishing associations between mental health disorders and educational attainment initially observed in Norway have been successfully replicated in England. Age at first diagnosis did not explain this trend, so future work should continue to explore possible explanations.

## Data availability statement

The data cannot be made publicly available, but can be accessed with permissions from both the Department for Education and South London and Maudsley NHS Foundation Trust. Supporting statistical code will become publicly available via AW’s GitHub account on publication: https://github.com/AliceWickersham.

## Ethics statement

CRIS has ethical approval from the South Central – Oxford C Research Ethics Committee (REC) as a database for secondary research (23/SC/0257, renewed 2023). The linkage between CRIS and the NPD has ethical approval from South Central – Oxford C Research Ethics Committee (25/SC/0229, renewed 2025).

## Patient consent statement

CRIS operates on an opt-out basis; individual consent/assent is not required for CRIS research. This project was approved by the CRIS Oversight Committee (project number 24-049). Section 251 support to process confidential patient information for linkage without consent was provided by the Health Research Authority Confidentiality Advisory Group, most recently in 2020 (20/CAG/0068).

## Acknowledgements

None.

## Abbreviations

ADHD: Attention Deficit Hyperactivity Disorder

## Key messages

- A Norwegian study showed weakening associations between pupil ADHD/internalising disorder diagnoses and end-of-school exam performance over a period of ten years.
- We replicated this finding in a large cohort drawn from secondary mental health services in South London, England.
- The trend was not explained by earlier age at first diagnosis.
- We also found a weakening association between pupil internalising disorder diagnosis and performance on exams taken at a younger age.
- While these relationships are yet to be fully explained, our findings highlight that growing availability of linked administrative data has made it feasible to replicate public health research internationally.

