## Supplement for "Mental health and educational attainment: Replicating diminishing associations in an England cohort"

*Table S1: STROBE Statement—Checklist of items that should be included in reports of cohort studies.*

| | Item No | Recommendation | Page # |
|---|---|---|---|
| **Title and abstract** | 1 | (*a*) Indicate the study's design with a commonly used term in the title or the abstract | 3 |
| | | (*b*) Provide in the abstract an informative and balanced summary of what was done and what was found | 3 |
| **Introduction** | | | |
| Background/rationale | 2 | Explain the scientific background and rationale for the investigation being reported | 4 |
| Objectives | 3 | State specific objectives, including any prespecified hypotheses | 4 |
| **Methods** | | | |
| Study design | 4 | Present key elements of study design early in the paper | 5 |
| Setting | 5 | Describe the setting, locations, and relevant dates, including periods of recruitment, exposure, follow-up, and data collection | 5 |
| Participants | 6 | (*a*) Give the eligibility criteria, and the sources and methods of selection of participants. Describe methods of follow-up | 5 |
| | | (*b*) For matched studies, give matching criteria and number of exposed and unexposed | 5 |
| Variables | 7 | Clearly define all outcomes, exposures, predictors, potential confounders, and effect modifiers. Give diagnostic criteria, if applicable | 5 |
| Data sources/ measurement | 8* | For each variable of interest, give sources of data and details of methods of assessment (measurement). Describe comparability of assessment methods if there is more than one group | 5 |
| Bias | 9 | Describe any efforts to address potential sources of bias | N/A |
| Study size | 10 | Explain how the study size was arrived at | 6 |
| Quantitative variables | 11 | Explain how quantitative variables were handled in the analyses. If applicable, describe which groupings were chosen and why | 5 |
| Statistical methods | 12 | (*a*) Describe all statistical methods, including those used to control for confounding | 5-6 |
| | | (*b*) Describe any methods used to examine subgroups and interactions | 6 |
| | | (*c*) Explain how missing data were addressed | 6 |
| | | (*d*) If applicable, explain how loss to follow-up was addressed | N/A |
| | | (*e*) Describe any sensitivity analyses | N/A |
| **Results** | | | |
| Participants | 13* | (a) Report numbers of individuals at each stage of study—eg numbers potentially eligible, examined for eligibility, confirmed eligible, included in the study, completing follow-up, and analysed | 5-6 |
| | | (b) Give reasons for non-participation at each stage | N/A |
| | | (c) Consider use of a flow diagram | N/A |

| | | | |
|---|---|---|---|
| Descriptive data | 14* | (a) Give characteristics of study participants (eg demographic, clinical, social) and information on exposures and potential confounders | 6 |
| | | (b) Indicate number of participants with missing data for each variable of interest | 6 |
| | | (c) Summarise follow-up time (eg, average and total amount) | 6 |
| Outcome data | 15* | Report numbers of outcome events or summary measures over time | 6 |
| Main results | 16 | (*a*) Give unadjusted estimates and, if applicable, confounder-adjusted estimates and their precision (eg, 95% confidence interval). Make clear which confounders were adjusted for and why they were included | 7-10 |
| | | (*b*) Report category boundaries when continuous variables were categorized | N/A |
| | | (*c*) If relevant, consider translating estimates of relative risk into absolute risk for a meaningful time period | N/A |
| Other analyses | 17 | Report other analyses done—eg analyses of subgroups and interactions, and sensitivity analyses | 9-10 |
| **Discussion** | | | |
| Key results | 18 | Summarise key results with reference to study objectives | 11 |
| Limitations | 19 | Discuss limitations of the study, taking into account sources of potential bias or imprecision. Discuss both direction and magnitude of any potential bias | 13 |
| Interpretation | 20 | Give a cautious overall interpretation of results considering objectives, limitations, multiplicity of analyses, results from similar studies, and other relevant evidence | 11-13 |
| Generalisability | 21 | Discuss the generalisability (external validity) of the study results | 13 |
| **Other information** | | | |
| Funding | 22 | Give the source of funding and the role of the funders for the present study and, if applicable, for the original study on which the present article is based | 1 |

*Give information separately for exposed and unexposed groups.

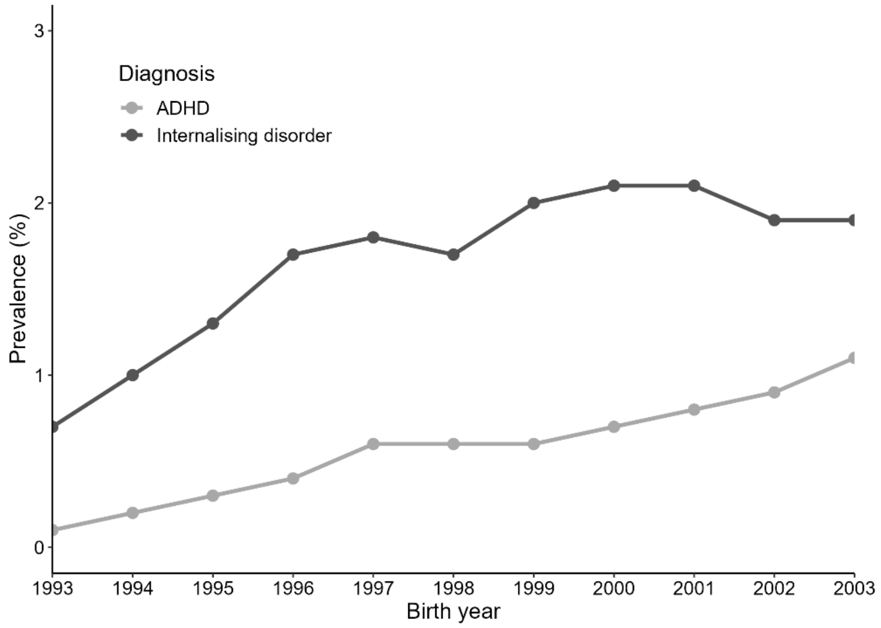


*Figure S1: Lifetime prevalence (birth – end of Year 11) of recorded mental health diagnosis for each birth cohort.*

*Table S1: Full regression results for ADHD diagnosis and Year 11 attainment, n=149,841.*

| Exposure | b coefficient (95% CI) | p-value |
|---|---|---|
| Intercept | -0.14 (-0.15 to -0.13) | <0.001 |
| ADHD diagnosis | -1.67 (-1.85 to -1.49) | <0.001 |
| Birth year | 0.00 (-0.00 to 0.00) | 0.901 |
| Gender (female) | 0.30 (0.29 to 0.31) | <0.001 |
| ADHD diagnosis x birth year interaction | 0.06 (0.03 to 0.08) | <0.001 |

*Table S2: Full regression results for internalising disorder diagnosis and Year 11 attainment, n=149,841.*

| Exposure | b coefficient (95% CI | p-value |
|---|---|---|
| Intercept | -0.15 (-0.16 to -0.13) | <0.001 |
| Internalising disorder diagnosis | -0.80 (-0.89 to -0.71) | <0.001 |
| Birth year | -0.00 (-0.00 to 0.00) | 0.322 |
| Gender (female) | 0.32 (0.31 to 0.33) | <0.001 |
| Internalising disorder diagnosis x birth year interaction | 0.05 (0.03 to 0.06) | <0.001 |

*Table S3: Cross-tabulation comparing pupils with and without ADHD diagnoses before Year 6 and Year 11 exams (n = 85,055).*

| | **No ADHD diagnosis before end of Year 11** | **ADHD diagnosis before end of Year 11** | **Total** |
|---|---|---|---|
| **No ADHD diagnosis before end of Year 6** | 84,427 | 370 | 84,797 |
| **ADHD diagnosis before end of Year 6** | Empty by definition | 258 | 258 |
| **Total** | 84,427 | 628* | 85,055 |

**Discrepancy with number reported in Year 11 main analysis is because not all pupils with Year 11 exam results had available Year 6 exam results.*

*Table S4: Cross-tabulation comparing pupils with and without internalising disorder diagnoses before Year 6 and Year 11 exams (n = 85,055).*

| | **No internalising disorder diagnosis before end of Year 11** | **Internalising disorder diagnosis before end of Year 11** | **Total** |
|---|---|---|---|
| **No internalising disorder diagnosis before end of Year 6** | 83,347 | 1,460 | 84,807 |
| **Internalising disorder diagnosis before end of Year 6** | Empty by definition | 248 | 248 |
| **Total** | 83,347 | 1,708* | 85,055 |

**Discrepancy with number reported in Year 11 main analysis is because not all pupils with Year 11 exam results had available Year 6 exam results.*

*Table S5: Full regression results for ADHD diagnosis and Year 6 attainment, n=85,055.*

| Exposure | b coefficient (95% CI) | p-value |
|---|---|---|
| Intercept | -0.05 (-0.06 to -0.03) | <0.001 |
| ADHD diagnosis | -1.09 (-1.37 to -0.81) | <0.001 |
| Birth year | -0.00 (-0.01 to 0.00) | 0.185 |
| Gender (female) | 0.10 (0.09 to 0.12) | <0.001 |
| ADHD diagnosis x birth year interaction | 0.05 (-0.02 to 0.11) | 0.166 |

*Table S6: Full regression results for internalising disorder diagnosis and Year 6 attainment, n=85,055.*

| Exposure | b coefficient (95% CI | p-value |
|---|---|---|
| Intercept | -0.05 (-0.06 to -0.04) | <0.001 |
| Internalising disorder diagnosis | -0.54 (-0.84 to -0.25) | <0.001 |
| Birth year | -0.00 (-0.01 to 0.00) | 0.086 |
| Gender (female) | 0.11 (0.09 to 0.12) | <0.001 |
| Internalising disorder diagnosis x birth year interaction | 0.10 (0.03 to 0.18) | 0.007 |
